# Durability of antibody responses and frequency of clinical and subclinical SARS-CoV-2 infection six months after BNT162b2 COVID-19 vaccination in healthcare workers

**DOI:** 10.1101/2021.10.16.21265087

**Authors:** Eric D. Laing, Carol D. Weiss, Emily C. Samuels, Si’Ana A. Coggins, Wei Wang, Richard Wang, Russell Vassell, Spencer L. Sterling, Marana A. Tso, Tonia Conner, Emilie Goguet, Matthew Moser, Belinda M. Jackson-Thompson, Luca Illinik, Julian Davies, Orlando Ortega, Edward Parmelee, Monique Hollis-Perry, Santina E. Maiolatesi, Gregory Wang, Kathleen F. Ramsey, Anatalio E. Reyes, Yolanda Alcorta, Mimi A. Wong, Alyssa R. Lindrose, Christopher A. Duplessis, David R. Tribble, Allison M.W. Malloy, Timothy H. Burgess, Simon D. Pollett, Cara H. Olsen, Christopher C. Broder, Edward Mitre

## Abstract

Antibodies against SARS-CoV-2 decay but persist six months post-vaccination, with lower levels of neutralizing titers against Delta than wild-type. Only 2 of 227 vaccinated healthcare workers experienced outpatient symptomatic breakthrough infections despite 59 of 227 exhibiting serological evidence of exposure to SARS-CoV-2 as defined by development of anti-nucleocapsid protein antibodies.

## Introduction

Neutralizing and binding antibodies appear to be associated with protection against symptomatic SARS-CoV-2 infection and COVID-19 (1, 2). Early assessments of the Pfizer-BioNTech BNT162b2 COVID-19 mRNA vaccine observed > 95% effectiveness against predominantly Alpha infections (3), but the potential impact of waning, post-vaccine neutralizing titers is an ongoing concern (4).

Apparent increases in vaccine-breakthrough infections may be due to waning antibody titers, increases in exposure risk as social distancing and mask wearing behaviors have decreased, and reduced vaccine effectiveness against the Delta variant. Since mid-2021, Delta has become the dominant genotype in the U.S. (5) and may cause increased hospitalization rates in unvaccinated individuals compared to other variants (6). Additionally, Delta has increased transmissibility compared to Alpha (7).

Because of growing concerns of vaccine-breakthrough risk, there has been a recent focus on vaccine boosting strategies in higher risk groups. However, uncertainty and debate remains about the need for vaccine boosters in generally healthy adults, including healthcare workers, because of limited post-vaccination immunological and clinical data (8).

Here, we report the binding antibodies (bAb) and neutralizing antibodies (nAb) as well as clinically overt and asymptomatic breakthrough infections that have occurred one year into the Prospective Assessment of SARS-CoV-2 Seroconversion (PASS) study (9).

### The Study

Generally healthy, adult HCWs at the Walter Reed National Military Medical Center (WRNMMC) who were seronegative for IgG antibodies to SARS-CoV-2 spike glycoprotein (spike) and had no history of COVID-19 were enrolled and followed in the PASS study as described in Jackson-Thompson *et al*. (9). Participants’ serum samples were collected monthly, diluted 1:400, and screened for immunoglobulin G (IgG) reactivity against SARS-CoV-2 spike and nucleocapsid protein (NP) in multiplex microsphere-based immunoassays, as previously described (See Supplement). Additionally, participants were asked to obtain nasopharyngeal SARS-CoV-2 PCR testing at the WRNMMC COVID-19 testing center upon experiencing symptoms consistent with SARS-CoV-2 infection.

Quantification of spike-specific IgG antibody levels in World Health Organization BAU was achieved by diluting serum samples at 1:400 and 1:8000, testing in a multiplex immunoassay composed of SARS-CoV-2 spike and NP coupled microspheres, and then interpolating against an internal standard curve calibrated to the NCI FNL U.S. SARS-CoV-2 IgG national standard (See Supplement, eFigure 1). Serum samples were assessed for nAb against SARS-CoV-2 WT and Delta as previously described using a well-characterized SARS-CoV-2 lentiviral pseudovirus neutralization assay to measure the potency of therapeutic antibodies against emerging variants (See Supplement). All data were log10-transformed. bAb were analyzed by Wilcoxon matched-pairs signed rank test and nAb by repeated measures Friedman ANOVA with Dunn’s multiple comparisons performed post-hoc using GraphPad Prism v9.0.

Excluding individuals infected prior to January 31 of 2021, the study followed 227 participants fully vaccinated with BNT162b2 and 17 unvaccinated participants. Participants were generally healthy, with a mean age of 41.7 years (range 20 - 69) and were predominantly female (Table 1).

**Table 1.**
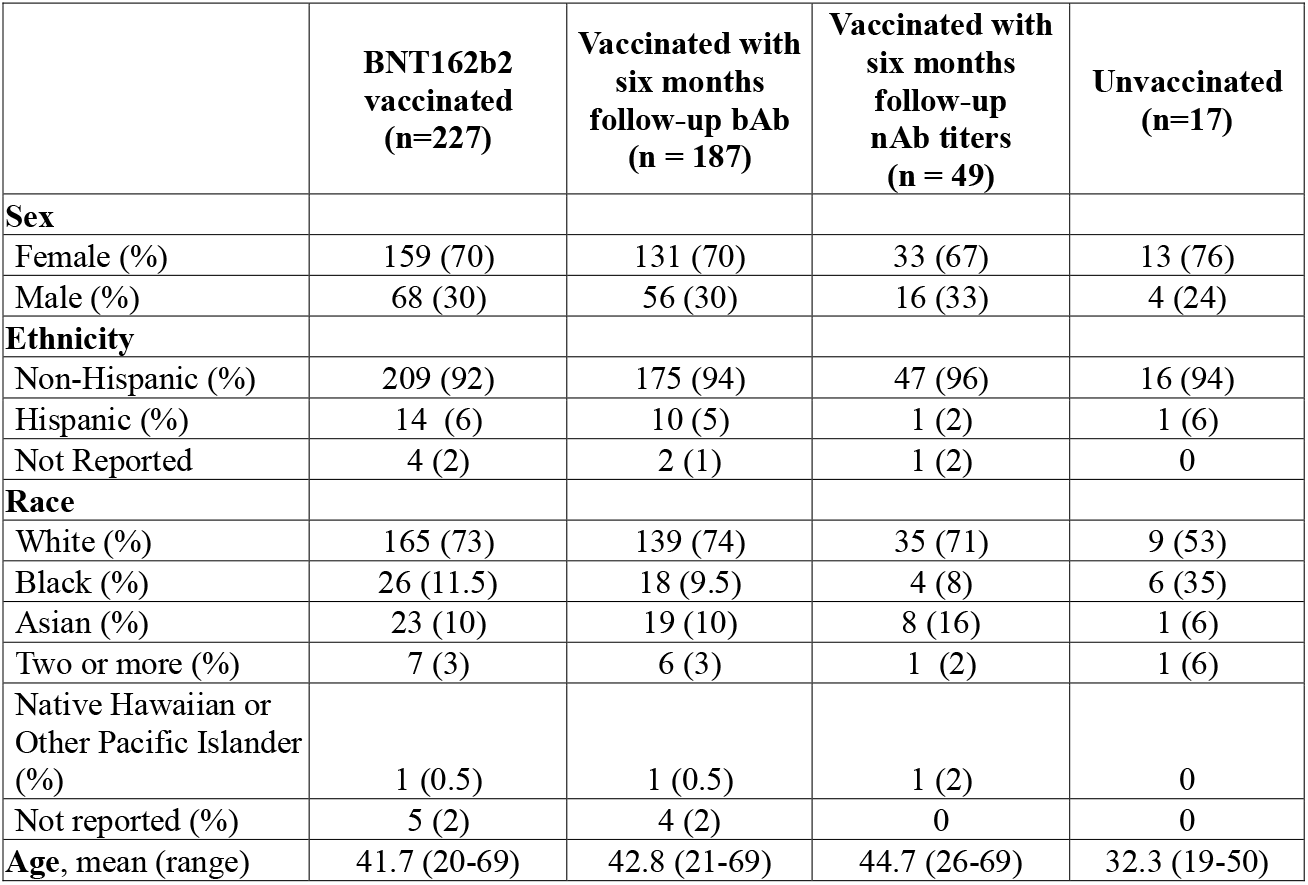
Participant demographics.

Seroconversion was observed in all participants one month following the second vaccine dose (Figure 1A). We quantified IgG bAb against spike at one and six months after full vaccination in the 187 vaccinated participants with serum samples at both time points. Anti-spike IgG bAb decreased from a geometric mean (GM) of 1929 BAU/mL (95%CI: 1752-2124) at one month post-vaccination to a GM of 442 BAU/mL (95%CI: 399-490) at six months post-vaccination (*P* < 0.001). Similarly, we observed decay of nAb between the one and six month-post vaccination time points. Peak SARS-CoV-2 WT nAb decreased from a geometric mean titer (GMT) of 551 (95% CI: 455-669) to 98 GMT (95% CI: 78-124) six months after full vaccination. The nAb GMTs were significantly higher against SARS-CoV-2 WT compared to SARS-CoV-2 Delta at both time points after vaccination (Figure 1C). In comparison, nAb against Delta decayed from 279 GMT (95% CI: 219-355) at peak to 38 GMT (95% CI: 31-48) after six months. Quantitative IgG bAb in BAU/ml correlated with nAb titers (⍰= 0.70, *P* < 0.001), demonstrating comparable decay of IgG bAb and nAb titers (eFigure 2).

**Figure 1.**
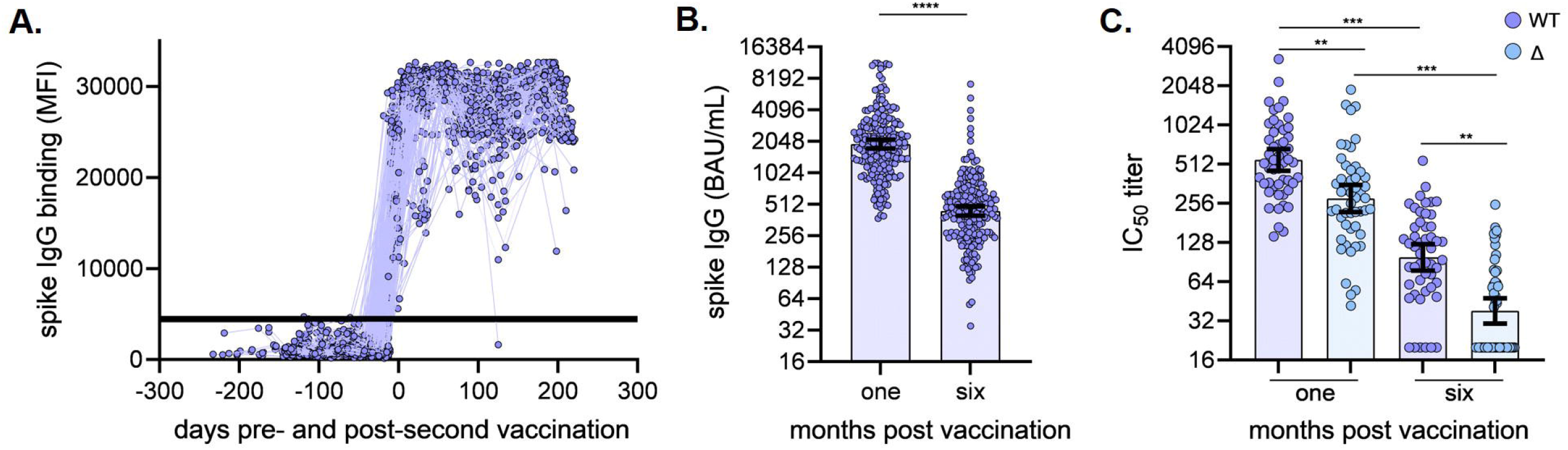
Vaccine-induced binding and neutralizing antibody responses. **(A)** Kinetics of vaccine-induced spike IgG bAb preceding and following second vaccination, participants n=227; MFI, median fluorescence intensity; shaded area indicates the positive/negative anti-spike IgG threshold. **(B)** Spike IgG bAb (BAU/mL) were quantified from serum samples collected one month (mean 36.9 days, range 23-81 days) and six months (mean 201.1 days, range 151-237 days) post vaccination, participants n=187; Wilcoxon matched-pairs signed rank test **** *P* < 0.001, two-tailed; y-axis is log2-scale **(C)** nAb titers against SARS-CoV-2 wild-type (WT) and Delta (Δ) variant from serum samples collected one month (mean 30.8 days, range 28-42 days) and six months (mean 200.1 days, range 189-219 days) post vaccination, participants n=49; Friedman ANOVA with Dunn’s multiple comparisons performed post-hoc *** *P* < 0.001, ** *P <* .01; y-axis is log2-scale. All errors bars represent the geometric mean and 95% confidence intervals.

In addition to anti-spike IgG bAb, we also monitored for seroconversion to IgG bAb to NP. Of vaccinated subjects, 26.0% (59/227) developed NP seroconversion between March and August of 2021 (Figure 2). Only two had symptomatic, PCR-positive vaccine breakthrough infections, both of which were self-limited, outpatient cases. In the unvaccinated cohort, four participants were diagnosed with SARS-CoV-2 infection: two by PCR when experiencing symptomatic infection (one outpatient case, one requiring inpatient ICU care) and two with pauci-symptomatic infection (both diagnosed by spike IgG seroconversion who reported mild symptoms retrospectively). The frequency of NP seroconversions in the vaccinated population correlated with the frequency of SARS-CoV-2 infections diagnosed in the unvaccinated participants, 23.5% (4/17) (Figure 2), suggesting similar exposure rates.

**Figure 2.**
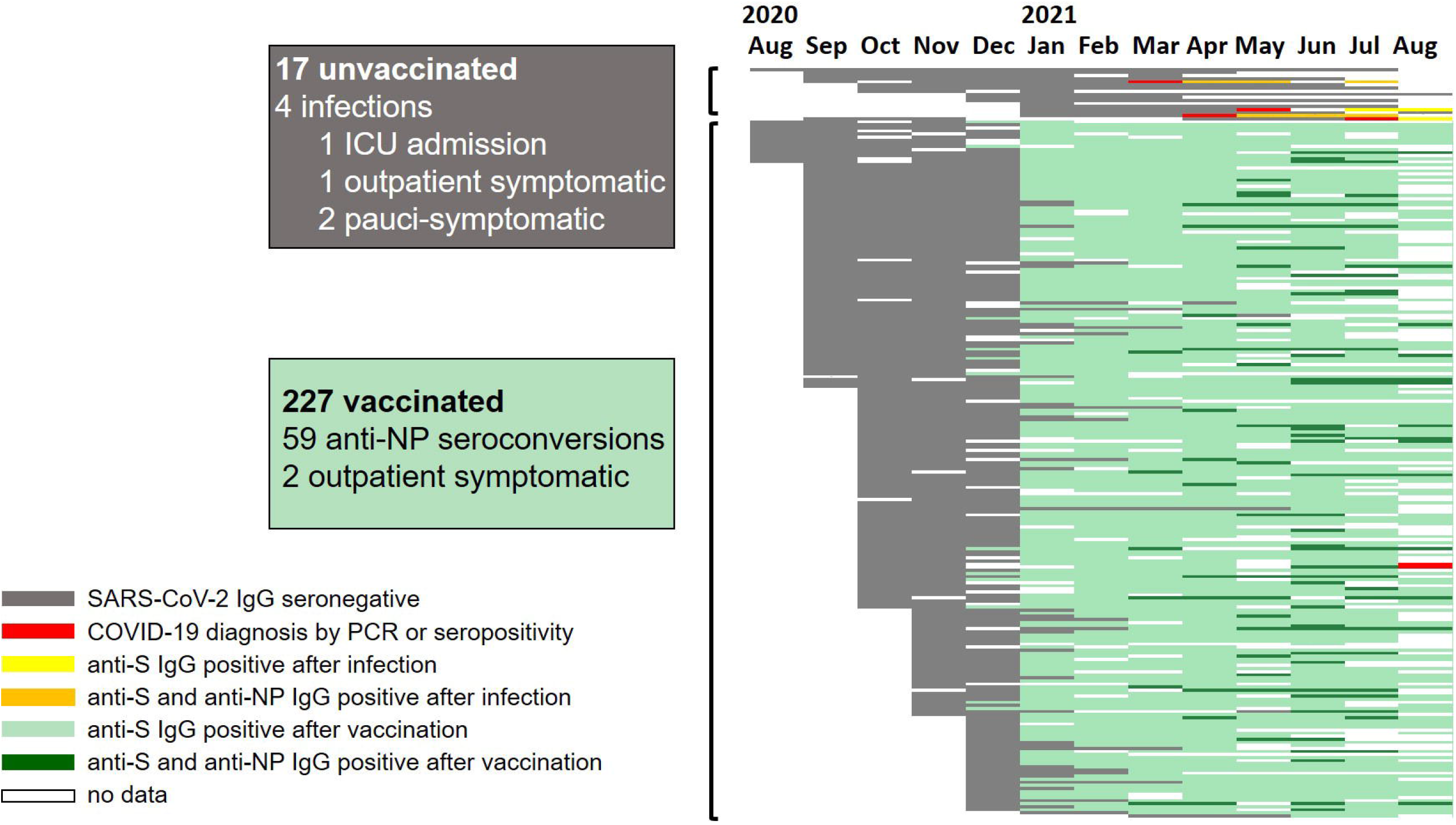
Timeline of antibody responses and SARS-CoV-2 infections in PASS participants. Each horizontal bar represents the infection, vaccination, and serological status obtained on a monthly basis of all PASS participants that had not been diagnosed with SARS-CoV-2 by PCR or anti-spike (S) IgG seroconversion by January 31, 2021. White spaces indicate no data. Grey bars represent negative anti-S IgG. Red bars indicate month of SARS-CoV-2 diagnosis by PCR positivity or anti-S IgG seroconversion. Yellow bars indicate anti-S IgG seroconversion after SARS-CoV-2 diagnosis in unvaccinated individuals, and orange bars indicate presence of both anti-S and anti-NP antibodies in unvaccinated individuals. Light green bars indicate anti-S IgG seroconversion after vaccination. Dark green bars indicate detection of anti-NP IgG antibodies in addition to anti-S antibodies at timepoints post-vaccination. N = 17 unvaccinated and 227 vaccinated participants.

## Conclusions

In this prospective cohort study of generally healthy adult HCWs, we find that SARS-CoV-2 spike IgG bAb and nAb induced by BNT162b2 mRNA COVID-19 vaccination wane, but persist through six months following vaccination, corroborating results of another study (10). Consistent with another report (11), significantly lower vaccine-induced nAb titers were observed against Delta compared to WT. Asymptomatic infections determined by NP seroconversions were relatively frequent, but symptomatic infection was rare, and severe disease was absent.

We observed one of 17 unvaccinated individuals develop severe COVID-19, versus no severe cases in 227 vaccinated participants. Of vaccinated individuals, there were two symptomatic, PCR-proven breakthrough infections, both of which were managed as outpatients. Interestingly, we observed that 26% of vaccinated participants developed antibodies against SARS-CoV-2 NP, suggesting that vaccinated subjects experienced exposures to SARS-CoV-2 as frequently as the unvaccinated population, yet rarely developed overt clinical disease. These results suggest that the BNT162b2 vaccine confers protection against severe clinical disease for at least six months in generally healthy adults, even in the face of frequent exposures to the virus and waning antibodies, though does not protect fully against mild disease.

The strengths of the study include the frequency of serological assessments and the use of variant specific nAb in addition to multiplexed antigen-specific IgG detection. Use of longitudinal serological assessments enabled detection of asymptomatic and/or pauci-symptomatic SARS-CoV-2 exposures, in addition to PCR testing triggered when participants exhibited symptoms. While we were powered to show clear differences in antibody titers over time, the study limitations include the moderate size of the cohort. Further, NP has lower sensitivity and specificity compared to spike protein, and is therefore a limited marker for subclinical SARS-CoV-2 infection (12). Indeed, NP seroconversion reportedly occurred in only 71% of PCR-confirmed vaccine breakthrough infections and is correlated with COVID-19 severity (13).

We observed persistence of nAb titers against SARS-CoV-2 WT equal to or greater than the lowest dilution tested in 90% (44/49) of healthy adults six months after vaccination with BNT162b2. Neutralizing activity against Delta was lower, with only 47% (23/49) of participants maintaining nAb titers above the lowest dilution at six months post-vaccination. The decrease in nAb does not necessarily mean that individuals have lost protection against severe COVID-19, as nAb titers required for protection are not known and neutralization is only one function of antibodies. In addition, memory B cells and T cells have been detected 8-12 months after SARS-CoV-2 infection, demonstrating that adaptive immune memory can be long-lasting to SARS-CoV-2 (14, 15). Further research is needed to understand the correlates of protection against moderate to severe COVID-19, and SARS-CoV-2 variants.

## Data Availability

All data produced in the present study are available upon reasonable request to the authors

## Funding

This work was supported by awards from the Defense Health Program and the CARES Act [HU00012020067] and the National Institute of Allergy and Infectious Disease [HU00011920111]. The protocol was executed by the Infectious Disease Clinical Research Program (IDCRP), a Department of Defense (DoD) program executed by the Uniformed Services University of the Health Sciences (USUHS) through a cooperative agreement by the Henry M. Jackson Foundation for the Advancement of Military Medicine, Inc. (HJF). This project has been funded in part by the National Institute of Allergy and Infectious Diseases at the National Institutes of Health, under an interagency agreement [Y1-AI-5072].

## Disclaimer

EDL, CDW, WW, RV, TC, KMH-P, CAD, AMWM, THB, CO, CCB, and EM are military Service members or employees of the U.S. Government. This work was prepared as part of their official duties. Title 17, U.S.C., §105 provides that copyright protection under this title is not available for any work of the U.S. Government. Title 17, U.S.C., §101 defines a U.S. Government work as a work prepared by a military Service member or employee of the U.S. Government as part of that person’s official duties. The views expressed in this article are those of the authors and do not necessarily reflect the official policy or position of the Uniformed Services University of the Health Sciences, Walter Reed National Military Medical Center, Naval Medical Research Center, the Department of Navy, the Department of Defense, the Henry. M. Jackson Foundation for the Advancement of Military Medicine, the U.S. Food and Drug Administration, or the U.S. Government. Mention of trade names, commercial products, or organizations does not imply endorsement by the U.S. Government.

## Potential Conflicts of Interest

DRT, THB, SDP: The Uniformed Services University (USU) Infectious Diseases Clinical Research Program (IDCRP), a U.S. Department of Defense institution, and the Henry M. Jackson Foundation (HJF) were funded under a Cooperative Research and Development Agreement to conduct an unrelated Phase III COVID-19 monoclonal antibody immune-prophylaxis trial sponsored by AstraZeneca. The HJF, in support of the USU IDCRP, was funded by the Department of Defense Joint Program Executive Office for Chemical, Biological, Radiological and Nuclear Defense to augment the conduct of an unrelated Phase III vaccine trial sponsored by AstraZeneca. Both these trials were part of the US Government COVID-19 response. Neither is related to the work presented here.

## Ethics Approval

This study protocol was approved by the Uniformed Services University of the Health Sciences Institutional Review Board (FWA 00001628; DoD Assurance P60001) in compliance with all applicable Federal regulations governing the protection of human participants. Written consent was obtained from all study participants.

## References

1. Khoury DS, Cromer D, Reynaldi A, Schlub TE, Wheatley AK, Juno JA, et al. Neutralizing antibody levels are highly predictive of immune protection from symptomatic SARS-CoV-2 infection. Nat Med. 2021 Jul;27(7):1205–11.

2. Gilbert PB, Montefiori DC, McDermott A, Fong Y, Benkeser DC, Deng W, et al. Immune Correlates Analysis of the mRNA-1273 COVID-19 Vaccine Efficacy Trial. medRxiv. 2021 Aug 15.

3. Thompson MG, Burgess JL, Naleway AL, Tyner H, Yoon SK, Meece J, et al. Prevention and Attenuation of Covid-19 with the BNT162b2 and mRNA-1273 Vaccines. N Engl J Med. 2021 Jul 22;385(4):320–9.

4. Fowlkes A, Gaglani M, Groover K, Thiese MS, Tyner H, Ellingson K. Effectiveness of COVID-19 Vaccines in Preventing SARS-CoV-2 Infection Among Frontline Workers Before and During B.1.617.2 (Delta) Variant Predominance - Eight U.S. Locations, December 2020-August 2021. MMWR Morb Mortal Wkly Rep. 2021 Aug 27;70(34):1167–9.

5. U.S. Centers for Disease Control and Prevention. COVID Data Tracker September 14, 2021 [cited September 15, 2021]; Available from: https://covid.cdc.gov/covid-data-tracker/#variant-proportions

6. Twohig KA, Nyberg T, Zaidi A, Thelwall S, Sinnathamby MA, Aliabadi S, et al. Hospital admission and emergency care attendance risk for SARS-CoV-2 delta (B.1.617.2) compared with alpha (B.1.1.7) variants of concern: a cohort study. Lancet Infect Dis. 2021 Aug 27.

7. Bolze A, Cirulli ET, Luo S, White S, Wyman D, Rossi AD, et al. SARS-CoV-2 variant Delta rapidly displaced variant Alpha in the United States and led to higher viral loads. medRxiv. 2021:2021.06.20.21259195.

8. Krause PR, Fleming TR, Peto R, Longini IM, Figueroa JP, Sterne JAC, et al. Considerations in boosting COVID-19 vaccine immune responses. The Lancet.

9. Jackson-Thompson BM, Goguet E, Laing ED, Olsen CH, Pollett S, Hollis-Perry KM, et al. Prospective Assessment of SARS-CoV-2 Seroconversion (PASS) study: an observational cohort study of SARS-CoV-2 infection and vaccination in healthcare workers. BMC Infect Dis. 2021 Jun 9;21(1):544.

10. Thomas SJ, Moreira ED, Kitchin N, Absalon J, Gurtman A, Lockhart S, et al. Six Month Safety and Efficacy of the BNT162b2 mRNA COVID-19 Vaccine. medRxiv. 2021:2021.07.28.21261159.

11. Chia PY, Xiang Ong SW, Chiew CJ, Ang LW, Chavatte J-M, Mak T-M, et al. Virological and serological kinetics of SARS-CoV-2 Delta variant vaccine-breakthrough infections: a multi-center cohort study. medRxiv. 2021:2021.07.28.21261295.

12. Laing ED, Sterling SL, Richard SA, Epsi NJ, Coggins S, Samuels EC, et al. Antigen-based multiplex strategies to discriminate SARS-CoV-2 natural and vaccine induced immunity from seasonal human coronavirus humoral responses. medRxiv. 2021 Feb 12.

13. Pollett SD, Richard SA, Fries AC, Simons MP, Mende K, Lalani T, et al. The SARS-CoV-2 mRNA vaccine breakthrough infection phenotype includes significant symptoms, live virus shedding, and viral genetic diversity. Clin Infect Dis. 2021 Jun 12.

14. Dan JM, Mateus J, Kato Y, Hastie KM, Yu ED, Faliti CE, et al. Immunological memory to SARS-CoV-2 assessed for up to 8 months after infection. Science. 2021;371(6529):eabf4063.

15. Turner JS, Kim W, Kalaidina E, Goss CW, Rauseo AM, Schmitz AJ, et al. SARS-CoV-2 infection induces long-lived bone marrow plasma cells in humans. Nature. 2021 2021/07/01;595(7867):421–5.

