## Supplementary Appendix for "Durability of antibody responses and frequency of clinical and subclinical SARS-CoV-2 infection six months after BNT162b2 COVID-19 vaccination in healthcare workers"

1. **eMethods**
2. **References**
3. **eFigures**
   1. **eFigure 1**
   2. **eFigure 2**

This supplemental material has been provided by the authors to give readers additional information about their work.

**1. eMethods**

**Multiplex microsphere-based immunoassay screening procedures**
Prefusion stabilized spike (S) glycoprotein ectodomain trimers (S-2P)^1,2^, hereafter referred to as spike, of SARS-CoV-2, HCoV-229E, and HCoV-NL63 were purchased from LakePharma, Inc (Hopkinton, MA, USA). HCoV-OC43 and HCoV-HKU1 spike were provided by Dr. Dominic Esposito, National Cancer Institute Frederick National Laboratory (NCI FNL), Protein Expression Laboratory, and have been previously described^3^. A SARS-CoV-2 NP was sourced from RayBiotech (Peachtree Corners, GA, USA). Multiplexed antigen-based antibody detection has been described previously ^4,5^. Briefly, SARS-CoV-2 spike and NP, and HCoV spike were coupled to magnetic microspheres (Bio-Rad, Hercules, CA, USA). Serum samples were collected from venipuncture in serum separator tubes, processed and stored at -80 °C in 500 µL aliquots until use. For weekly screening, neat human serum samples (1.25 µL) were diluted 1:400 in 1X PBS and heat inactivated at 60 ˚C for 30 min after dilutions. Diluted serum samples were incubated with a master mix of SARS-CoV-2, HCoV-OC43, HCoV-HKU1, HCoV-229E, HCoV-NL63 spike, and SARS-CoV-NP coupled microspheres. This multiplex microsphere-based immunoassay has a sensitivity to detect SARS-CoV-2 spike reactive IgG seroconversion between 7-28 days post-symptom onset of 94%, with specificity of 100%^4,6,7^.
After a 45 minute incubation of diluted serum and antigen-coupled microspheres, with agitation (900 rpm), plates were washed with PBS-Tween20 (0.05%) and 100 µL of biotinylated cross-absorbed anti-human IgG (Thermo Fisher Scientific, Waltham, MA) diluted in 1X PBS-T (1:5000) was added to each well, and plates were incubated for 45 minutes with agitation. Lastly, after washing, streptavidin-phycoerythrin was diluted 1:1000 in PBS-T, and 100 µL were added to each well and plates were incubated for 45 min with agitation (900 rpm). Plates were washed, and microspheres were resuspended with 100 µL PBS-T per well then analyzed on Bio-Plex 200 multiplexing systems (Bio-Rad) and median fluorescence intensity (MFI) values for samples are reported as the PBS adjusted average from duplicate plates. Antibody testing was blind to descriptive data, including demographic data, SARS-CoV-2 PCR status, and clinical phenotype.

**Calibration to NCI FNL U.S. serology standard and interpolation of binding antibody units (BAU/mL)**	An internal reference standard (IR-std), a mixture of nine PASS study serum samples obtained one month after PCR-confirmed SARS-CoV-2 infection in 2020, was calibrated against the NCI FNL U.S. serology national standard (US-std) for SARS-CoV-2 spike protein and NP IgG. The IR-std and US-std were diluted 2-fold starting at 1:400 through 1:512,000, and IgG was detected as described above. The concentration of spike-specific IgG in IR-std was determined to be 428 BAU/ml by averaging the results of four separate analyses interpolating IR-std against a standard curve of the US-std with known concentration of 764 BAU/ml. With the established IR-std BAU/mL, PASS participant serum samples were tested at 1:400 and 1:8000 dilutions. All MFI values were adjusted to the PBS-blank control wells, then MFI values were interpolated against the IR-std included on each 96-well microtiter plate.

**SARS-CoV-2 S-pseudovirus production and neutralization**	A codon-optimized spike gene corresponding to the Wuhan-1 spike with the D614G substitution was used to make the wild-type (WT) pseudovirus. A codon-optimized spike gene used to make the Delta (B.1.617.2) pseudovirus had the following mutations on the WT backbone: T19R, G142D, E156 deletion, F157 deletion, R158G, L452R, T478K, D614G, P681R, and D950N. This neutralization assay has been previously described^8,9^, briefly, 5μg of pCMVΔR8.2, 5μg of pHR’CMVLuc and 0.5μg of S expression plasmids were co-transfected in 293T cells. Pseudovirus supernatants were collected approximately 48 hours post-transfection, filtered through a 0.45 μm low protein binding filter, and used immediately or stored at -80^o^C. Pseudovirus titers were measured by infecting 293T-ACE2.TMPRSS2 cells, which stably express human angiotensin converting enzyme 2 (ACE2) and transmembrane serine protease 2 (TMPRSS2), for 48 hours prior to measuring luciferase activity (luciferase assay reagent, Promega, Madison, WI). Neutralization titers were calculated using a nonlinear regression curve fit (GraphPad Prism software Inc., La Jolla, CA) using 8-point dilution curves. The mean titer from at least two independent experiments each with intra-assay duplicates was reported as the final titer. Titers below the lowest serum dilution of 1:40 were treated as 20 for statistical analysis.

**2. References**

1.	Wrapp D, Wang N, Corbett KS, et al. Cryo-EM structure of the 2019-nCoV spike in the prefusion conformation. *Science.* 2020;367(6483):1260-1263.

2.	Esposito D, Mehalko J, Drew M, et al. Optimizing high-yield production of SARS-CoV-2 soluble spike trimers for serology assays. *Protein Expr Purif.* 2020;174:105686.

3.	Hicks J, Klumpp-Thomas C, Kalish H, et al. Serologic Cross-Reactivity of SARS-CoV-2 with Endemic and Seasonal Betacoronaviruses. *J Clin Immunol.* 2021;41(5):906-913.

4.	Laing ED, Sterling SL, Richard SA, et al. Antigen-based multiplex strategies to discriminate SARS-CoV-2 natural and vaccine induced immunity from seasonal human coronavirus humoral responses. *medRxiv.* 2021.

5.	Laing ED, Epsi NJ, Richard SA, et al. SARS-CoV-2 antibodies remain detectable 12 months after infection and antibody magnitude is associated with age and COVID-19 severity. *medRxiv.* 2021:2021.2004.2027.21256207.

6.	Clifton GT, Pati R, Krammer F, et al. SARS-CoV-2 Infection Risk Among Active Duty Military Members Deployed to a Field Hospital - New York City, April 2020. *MMWR Morb Mortal Wkly Rep.* 2021;70(9):308-311.

7.	Ramos I, Goforth C, Soares-Schanoski A, et al. Antibody Responses to SARS-CoV-2 Following an Outbreak Among Marine Recruits With Asymptomatic or Mild Infection. *Front Immunol.* 2021;12:681586.

8.	Lusvarghi S, Wang W, Herrup R, et al. Key substitutions in the spike protein of SARS-CoV-2 variants can predict resistance to monoclonal antibodies, but other substitutions can modify the effects. *bioRxiv.* 2021:2021.2007.2016.452748.

9.	Neerukonda SN, Vassell R, Herrup R, et al. Establishment of a well-characterized SARS-CoV-2 lentiviral pseudovirus neutralization assay using 293T cells with stable expression of ACE2 and TMPRSS2. *PLoS One.* 2021;16(3):e0248348.

**3. eFigures**

**eFigure 1. Comparison of the PASS internal reference standard (IR-std) to the NCI FNL U.S. serology national standard (US-std).**

A. **B.**

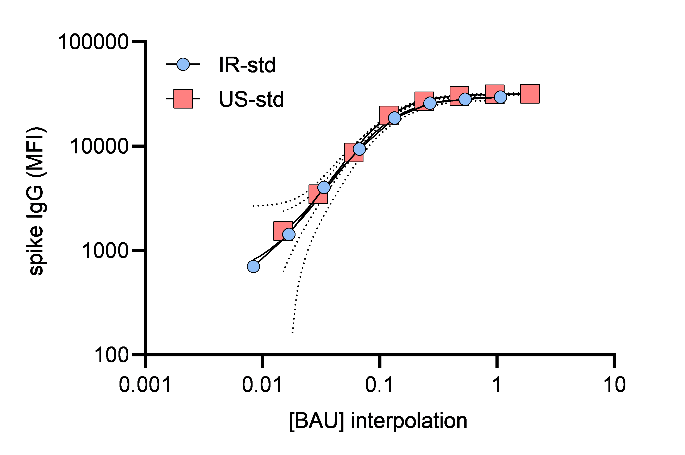

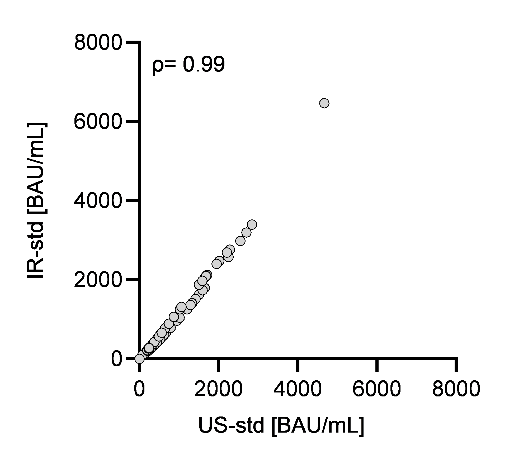

**A)** Comparison of the PASS internal reference standard (IR-std) curve to the NCI FNL U.S. serology national standard (US-std) curve for SARS-CoV-2 spike protein reactive immunoglobulin G (IgG); MFI, median fluorescence intensity; BAU, binding antibody units; curves with dashed lines represent the mean and error bars of independent experiments, axes are log10-scale,representative of four independent experiments. **B)** Correlation between spike IgG BAU/mL interpolated from the IR-std or the US-std; n=76 serum samples, Spearman’s rho (ρ) = 0.99, two-tailed *P* < 0.001.

**eFigure 2. Correlation between spike IgG bAb and nAb titers against SARS-CoV-2 WT six months-post vaccination.**

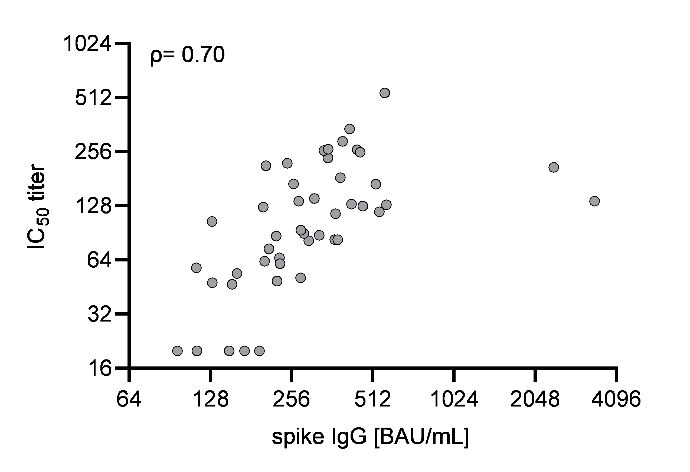

Six months post-vaccination serum antibodies were evaluated for correlation between spike IgG bAb, and nAb IC_50_ titers against SARS-CoV-2 WT, n = 49. Spearman’s rho (ρ) = 0.70, two-tailed *P* <0.001; axes are log2-scale.
